# MTXPK.org: A clinical decision support tool evaluating high-dose methotrexate pharmacokinetics to inform post-infusion care and use of glucarpidase

**DOI:** 10.1101/2020.04.08.20056713

**Authors:** Zachary L. Taylor, Tomoyuki Mizuno, Nieko C. Punt, Balaji Baskaran, Adriana Navarro Sainz, William Shuman, Nicholas Felicelli, Alexander A. Vinks, Jesper Heldrup, Laura B. Ramsey

**Affiliations:** Department of Molecular, Cellular, and Biochemical Pharmacology, University of Cincinnati, Cincinnati, OH; Division of Research in Patient Services, Cincinnati Children’s Hospital Medical Center, Cincinnati, OH; Division of Clinical Pharmacology, Cincinnati Children’s Hospital Medical Center, Cincinnati, OH; Department of Pediatrics, University of Cincinnati, Cincinnati, OH; Medimatics, Maastricht, The Netherlands; Division of Biomedical Informatics, Cincinnati Children’s Hospital Medical Center, Cincinnati, OH; Department of Pediatrics Oncology, Lund University Hospital, Lund, Sweden

**Keywords:** Bayesian, Chemotherapy, Modeling, NONMEM, Oncology, Pediatric, Population Pharmacokinetics, Software, Therapeutic Drug Monitoring, Translational pharmacokinetics-pharmacodynamics

## Abstract

Methotrexate (MTX), an anti-folate, is administered at high-doses to treat malignancies in children and adults. However, there is considerable interpatient variability in clearance of high-dose (HD) MTX. Patients with delayed clearance are at an increased risk for severe nephrotoxicity and life-threatening systemic MTX exposure. Glucarpidase is a rescue agent for severe MTX toxicity that reduces plasma MTX levels via hydrolysis of MTX into inactive metabolites, but is only indicated when MTX concentrations are > 2 standard deviations above the mean excretion curve specific for the given dose together with a significant creatinine increase (> 50%). Appropriate administration of glucarpidase is challenging due to the ambiguity in the labeled indication. A recent consensus guideline was published with an algorithm to provide clarity in when to administer glucarpidase, yet clinical interpretation of lab results that do not directly correspond to the algorithm prove to be a limitation of its use.

The goal of our study was to develop a clinical decision support tool to optimize the administration of glucarpidase for patients receiving HD MTX. Here, we describe the development of a novel three-compartment MTX population PK model using 31,672 MTX plasma concentrations from 772 pediatric patients receiving HD MTX for the treatment of acute lymphoblastic leukemia and its integration into the online clinical decision support tool, MTXPK.org. This web-based tool has the functionality to utilize individualized demographics, serum creatinine, and real-time drug concentrations to predict the elimination profile and facilitate model-informed administration of glucarpidase.

## Introduction

Methotrexate (MTX) is a commonly used anti-folate administered intravenously at high-doses to effectively treat pediatric and adult patients with acute lymphoblastic leukemia (ALL), osteosarcoma (OS), and lymphoma (1–4). Unfortunately, patients receiving intravenous (IV) high-dose (HD) MTX often experience significant toxicities of the kidney and eventually liver, and gastrointestinal tract. To reduce these toxicities, supportive care (including folate supplementation, fluid hyper hydration, and urine alkalization before and during the MTX treatment) is used to facilitate renal elimination (5). However, there is still considerable interpatient variability in clearance of HD MTX with severe delayed MTX clearance seen in 0.5-1% of pediatric ALL patients (6, 7), 1.8% of courses of 12 g/m^2^ over 4 hours in youth with OS (4), and 1-12% of adults treated for lymphoma (8, 9). Patients with delayed MTX clearance are at an increased risk for severe nephrotoxicity, which further reduces MTX elimination and can culminate in life-threatening systemic MTX exposure.

Currently, there is a lack of clinical tools for the identification of patients likely to experience delayed MTX clearance prior to the initiation of treatment or early after the infusion. As a result, response to patients with delayed MTX clearance tends to be reactive. Once concentrations rise above critical limits, rescue by folinic acid can be inadequate (7). In such cases, glucarpidase, an FDA-approved exogenous enzyme, is administered as a rescue agent that rapidly hydrolyzes MTX into two inactive metabolites that are eliminated via renal and non-renal pathways (6, 10–13). The rapid hydrolysis by glucarpidase reduces plasma MTX concentrations by > 90% within 15 minutes of administration (4, 14–17). Glucarpidase is only indicated when MTX concentrations are > 2 standard deviations (SD) above the mean excretion curve specific for the given dose (10) in order to avoid under-exposure to MTX and risk of relapse. Appropriate interpretation of the indication and administration of glucarpidase remains a challenge since many clinicians do not know the expected excretion curve and 2 times SD. As a result, a recent glucarpidase consensus guideline was published in an attempt to clarify the interpretation of the glucarpidase indication by providing a glucarpidase treatment algorithm that detailed MTX concentrations at select time points following a MTX infusion of several common dosing regimens (11). Although beneficial, the clinical interpretation of MTX concentrations that do not correspond to the algorithm’s time points prove to be a limitation of its use.

Therefore, the purpose of this study was to develop a web-based clinical decision support tool that provides a MTX population PK model-informed interpretation in tandem with the glucarpidase consensus guideline to facilitate the administration of glucarpidase in all patients receiving HD MTX, regardless of indication or age. Our aim was to develop a novel MTX population PK model using PK data from the Nordic Society for Pediatric Hematology and Oncology (NOPHO) and evaluate it as a model for the web-based clinical decision support tool, MTXPK.org. Although several PK models for HD MTX have been described, they may not describe patients with delayed clearance well because they are constructed from MTX concentrations only up to 44 hours after the start of MTX infusion (18–23). We hypothesize that our NOPHO PK model will better fit patients with delayed MTX clearance than currently available population PK models due to the dense PK sampling from patients with delayed MTX clearance with concentrations recorded up to 300 hours after the start of infusion.

## Methods

### Study Design

This study retrospectively analyzed PK data from 820 de-identified pediatric patients who were receiving HD MTX for the treatment of Philadelphia chromosome negative ALL between January 2002 and December 2014. Included in the current study were children (ages 1-18.83 years at diagnosis) from hospitals across Denmark, Finland, Lithuania, Norway, and Sweden receiving treatment in accordance with the NOPHO ALL2000 and ALL2008 protocols (2, 7, 24). Patients were not included if they received glucarpidase because the immunoassay measurements of MTX have interference from the DAMPA that results after MTX is cleaved by glucarpidase (11). Forty-eight patients and 679 concentrations were excluded from the PK analysis due to missing dosing information. In brief, patients were scheduled to receive 6-8 courses of intravenous HD MTX (5 or 8 g/m^2^) over a 24-hour infusion with folinic acid rescue occurring at 36 or 42 hours after the start of infusion. Serial plasma MTX levels were collected at 24, 36, and every 6 hours after the start of infusion until the plasma concentrations reached ≤ 0.2 µmol/L (7). MTX concentrations in plasma were quantified using one of two comparable immunoassay methods: enzyme-multiplied immunoassay technique (EMIT) or fluorescence polarization immunoassay (FPIA) (7, 25). Clinical covariates, like serum creatinine (SCr) levels were recorded prior to the start of each MTX infusion and at least daily in tandem with plasma MTX levels. Demographic covariates such as age, sex, body surface area (BSA), weight, and country of treatment were also included in the database. The NOPHO ALL2000 and ALL2008 protocols were approved by the Ethical Committee of the Capital Region of Denmark and by local ethical review boards.

### Model Development and Bayesian Estimation

We performed a population PK analysis of HD MTX using nonlinear mixed-effects modeling implemented in NONMEM^®^ version 7.2.0 (ICON, Ellicott City, MD) and validated using KinPop++. Two- and three-compartment structural models were considered. PK data was assumed to be log-normally distributed (26). The three-compartment with an exponential residual error model was chosen as the base model after considering objective function value (OFV) and goodness-of-fit (GOF) plots. Clinical (SCr) and demographic (BSA, weight, age, sex) data collected during the course of the MTX treatment were included in the covariate analysis. Additional information regarding the development of the population PK model can be found in the **Supplemental Methods**. A module for Bayesian estimation was included in the MTXPK.org platform using a three-compartment structure model developed using the visual PKPD model designer Edsim++ (Mediware, Prague, Czech Republic) and executed using an Edsim++ compatible PKPD modeling engine (27, 28).

### Evaluation of MTX Population PK Models

Two published MTX population PK models (18, 29) and our three-compartment MTX model were tested in an MTXPK.org prototype (MTXSim), which uses the same platform libraries as MTXPK.org. Model parameters and rationale for selection are detailed in the **Table S1; Supplementary Materials** (18, 29). These models were evaluated on their prediction bias and prediction precision using a small validation cohort of pediatric and young adult patients with ALL, lymphoma, and OS and whose data were not part of the modeling set (30).

### Integration into a Web-based Clinical Decision Support Tool

At the top level is the web application (**Figure S1, Supplementary Materials**). It was created so that it could run on both large and small screen devices by utilizing responsive web design. The application presents different design elements based on the user’s screen size and platform. It was built with React, a JavaScript library, and uses a Web API, to send data to, and receive data from a library containing all MTX specific business logic (BSA calculator and dosing schedules for each indication). This MTX specific library uses a general purpose PKPD modeling engine for simulation and Bayesian estimation, which in turn employs models from a PKPD model repository (28). MTX models were designed, executed, and validated using Edsim++ 1.80 (Mediware, Prague, Czech Republic) (27). The PKPD modeling engine runtime is capable of executing these models outside Edsim++. A stand-alone windows application (MtxSim) was used for end-to-end validation of the system (including the models) and as a prototype tool for the web development team building MTXPK.org. In MtxSim different models can be selected for testing.

### Statistics

Comparative statistical analysis between sex and country included the Student’s t test and one-way ANOVA, respectively (mean ± SD). A *p* < 0.05 was considered statistically significant. Linear regression was used to determine the R^2^ for the GOF plots. Statistical analysis was completed using GraphPad Prism 8.0.1.

## Results

### Population PK model development

The final population PK model was based on data from 772 patients, covering 4,986 courses, and 31,672 plasma MTX concentrations (**Figure 1**) – with 5,535 concentrations recorded ≥ 96 hours after the start of infusion. The demographic and PK information for the NOPHO dataset is summarized in **Table 1**. A three-compartment model using an exponential residual error model best described the PK of HD MTX in Nordic pediatric patients with ALL (**Figure 2**). The objective function value (OFV) for the base three-compartment model was significantly lower than that for the base two-compartment model (ΔOFV = −6,136.43, p < 0.001), suggesting that the three-compartment model provided significantly improved description of the data (31). The three-compartment model also demonstrated significant improvement in the population prediction performance (**Figure 2A**) compared to the two-compartment model (**Figure 2B**, p < 0.001), the latter which was prone to substantial model-based over-estimation of low MTX concentrations and MTX clearance. The three-compartment model demonstrated less error as the time after the dose increased (**Figure 2C**) compared to the two-compartment model (**Figure 2D**). Additional analysis of the residuals using the root mean square error (RMSE) as a measure of model accuracy revealed a 44% reduction in the unexplained variance while using the three-compartment model, further demonstrating that the three-compartment structural model out-performs the two-compartment model.

**Figure 1:**
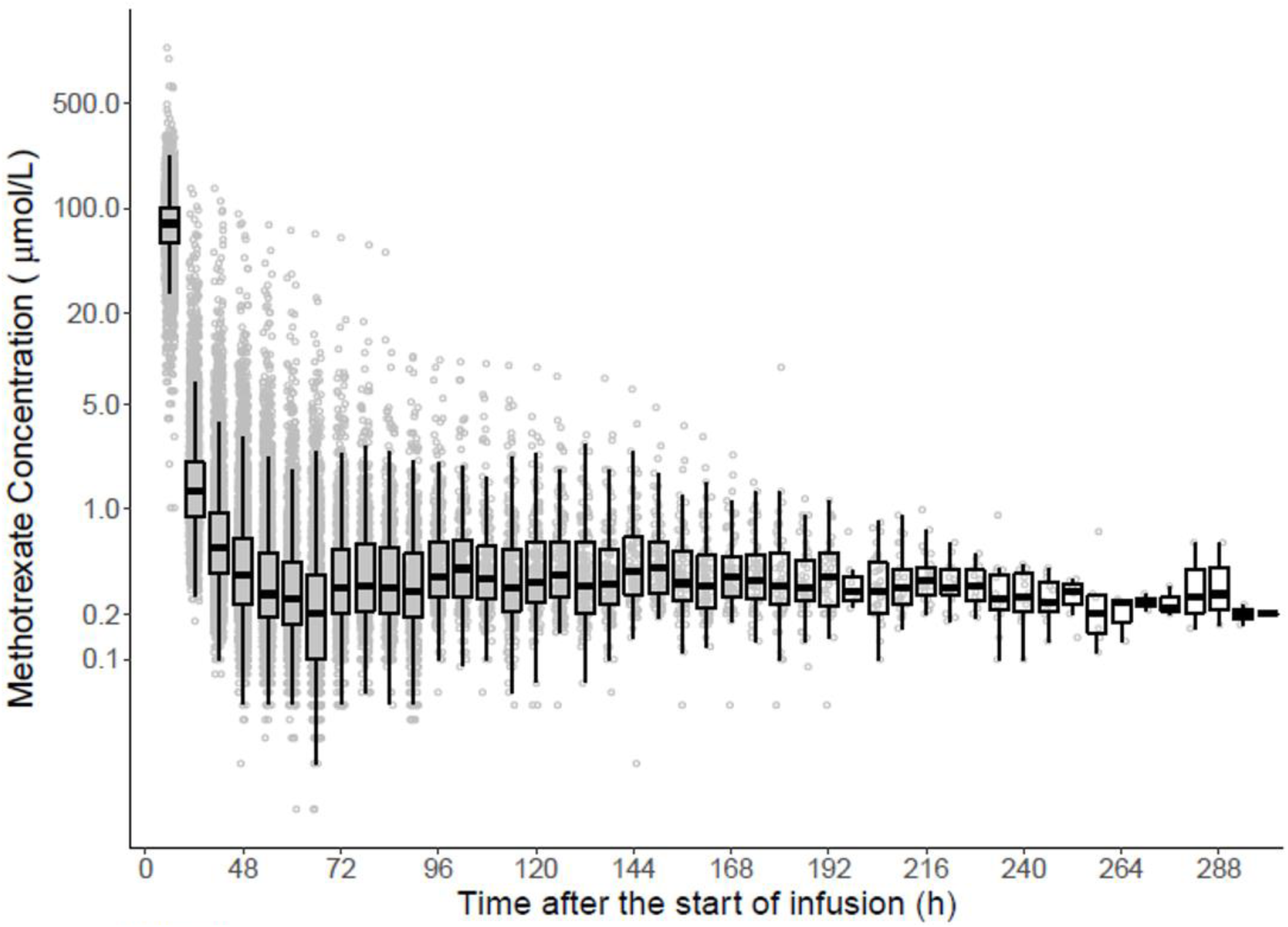
Concentration-time plot of the NOPHO dataset. Individual concentrations are in blue with a box and whiskers plot shown on top. The black line represents the median value. The box plot illustrates the inter-quartiles with the whiskers representing the 2 standard deviations.

**Table 1.** Patient demographics

|  | **Median (n)** | **Range (%)** |
| --- | --- | --- |
| **Age (Years)** | 4 | (1-18.83) |
| **Body Surface Area (m<sup>2</sup>)** | 0.745 | (0.4-2.31) |
| **Courses per Patient** | 8 | (1-8) |
| **Country** |  |  |
| Denmark | 207 | (27%) |
| Finland | 78 | (10%) |
| Norway | 59 | (8%) |
| Sweden | 428 | (55%) |
| **Dose (g/m<sup>2</sup>)** | 5 | (0.6-10.1) |
| **Sex (Female)** | 333 | (43%) |
| **Serum Creatinine (μmol/L)** | 29 | (4-155) |
| **Weight (kg)** | 17.8 | (7.2-105) |

n: Number of patients
m<sup>2</sup>: Meters squared
g/m<sup>2</sup>: Grams/meters squared
μmol/L: Micromole per liter
kg: Kilograms

**Figure 2:**
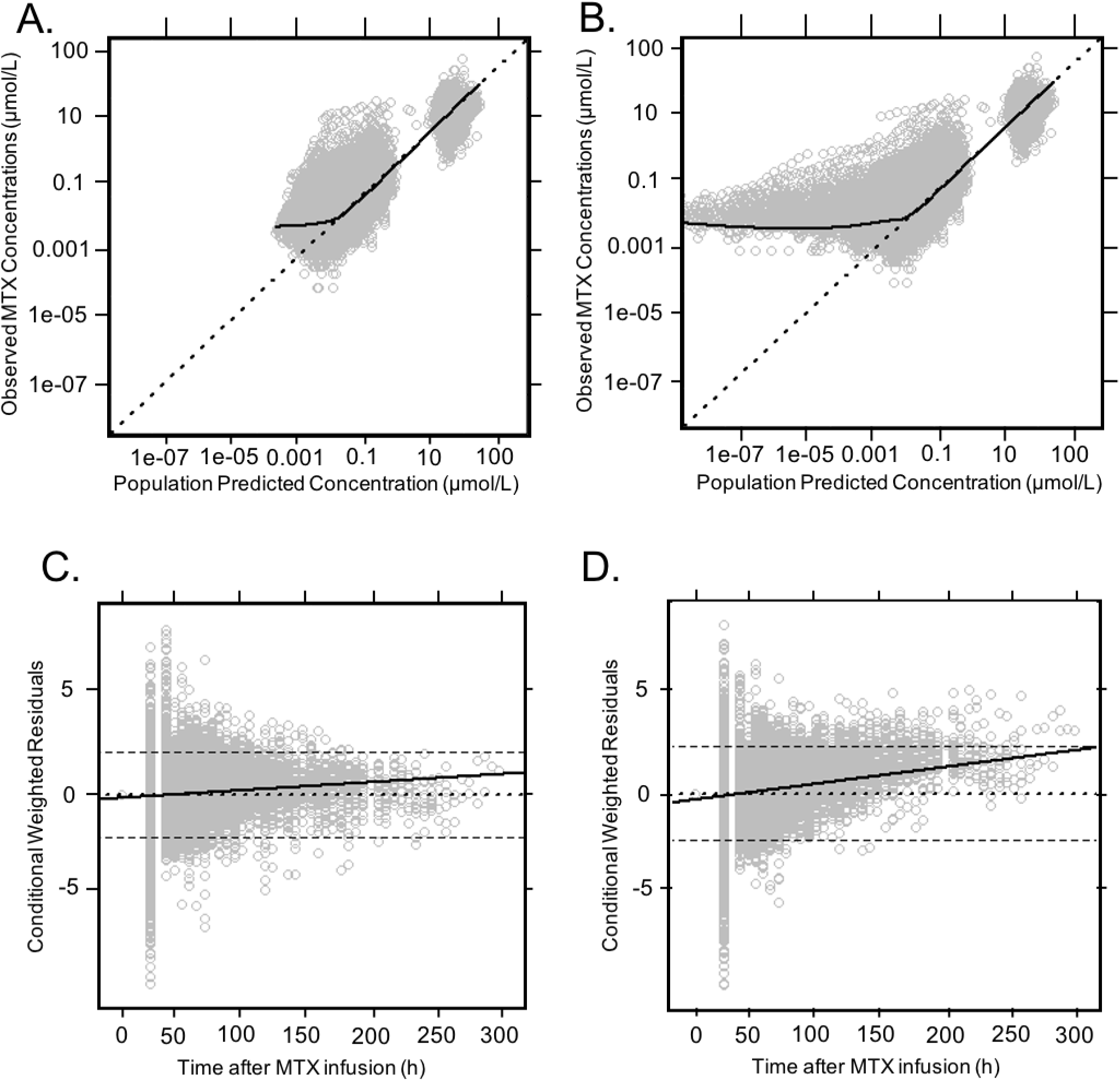
Comparing the GOF plots for two- and three-compartment structural models. The three-compartment model showed significantly less bias at lower predicted concentrations (A) compared to the two-compartment model (B), which displayed over-estimation of clearance. The three-compartment model displayed improved predictive performance at later times following a MTX infusion (C) compared to the two-compartment model (D). For figures A and B, the dotted line represents the line of identity. For figures C and D, the dotted line represents the zero-line. The dashed lines represent the 2 standard deviations. The solid black line represents the trend line for all figures.

### Covariate Model

The stepwise inclusion of covariates is presented in **Table 2**. Among the clinical and demographic covariates considered, the patient’s BSA yielded the largest reduction in OFV (ΔOFV = −2,358.81, p < 0.001) and displayed a coefficient of determination with clearance (**Figure 3A**, R^2^ = 0.54) than the patient’s body weight (**Figure 3B**, R^2^ = 0.36). All PK parameters (CL, V1, Q2, V2, Q3, and V3) were normalized to a BSA of 1.73 m^2^. After the clearance was normalized, demographic covariates such as age and sex did not significantly improve clearance estimates (**Figure S2**, **Supplementary Materials**). Finnish patients had an estimated 26% faster clearance compared to Swedish, Danish, and Norwegian patients (**Figure S3, Supplementary Materials**). SCr was negatively correlated to normalized clearance (**Figure 3C**), which was accounted for by incorporating SCr as a time-varying covariate into a power component normalized to the population median of 29 µmol/L (0.33 mg/dL). Once adjusted, SCr further explained inter-individual variability in clearance and reduced the OFV by −702.73 (p < 0.001). The equation for the final covariate model is presented below:

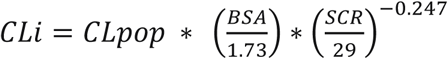

**Table 2:** Forward stepwise inclusion of covariates

| No. | Covariates | Parameter | Covariate Model | ΔOFV | P < 0.05 | Reference Model | Notes |
| --- | --- | --- | --- | --- | --- | --- | --- |
| 1 | Base Model | --- | --- | --- | --- | --- | 3-CP, IV, linear elimination |
| 2 | BSA | Cl | Normalize | -2358.81 | Yes | 1 | Normalized to 1.73 m<sup>2</sup> |
| 3 | Weight | Cl | Normalize | -1260.02 | Yes | 1 | Normalized to 70 kg |
| 4 | Serum Creatinine (SCR) | Cl | Power | -39.59 | Yes | 1 | (SCR/29)<sup>power</sup> |
| 5 | BSA + SCR | Cl | Power | -702.73 | Yes | 2 | Normalization + (SCR/29)<sup>power</sup> |

No.: Model number
ΔOFV: Change in the objective function value
CP: Compartment
IV: Intravenous
BSA: Body surface area

**Figure 3:**
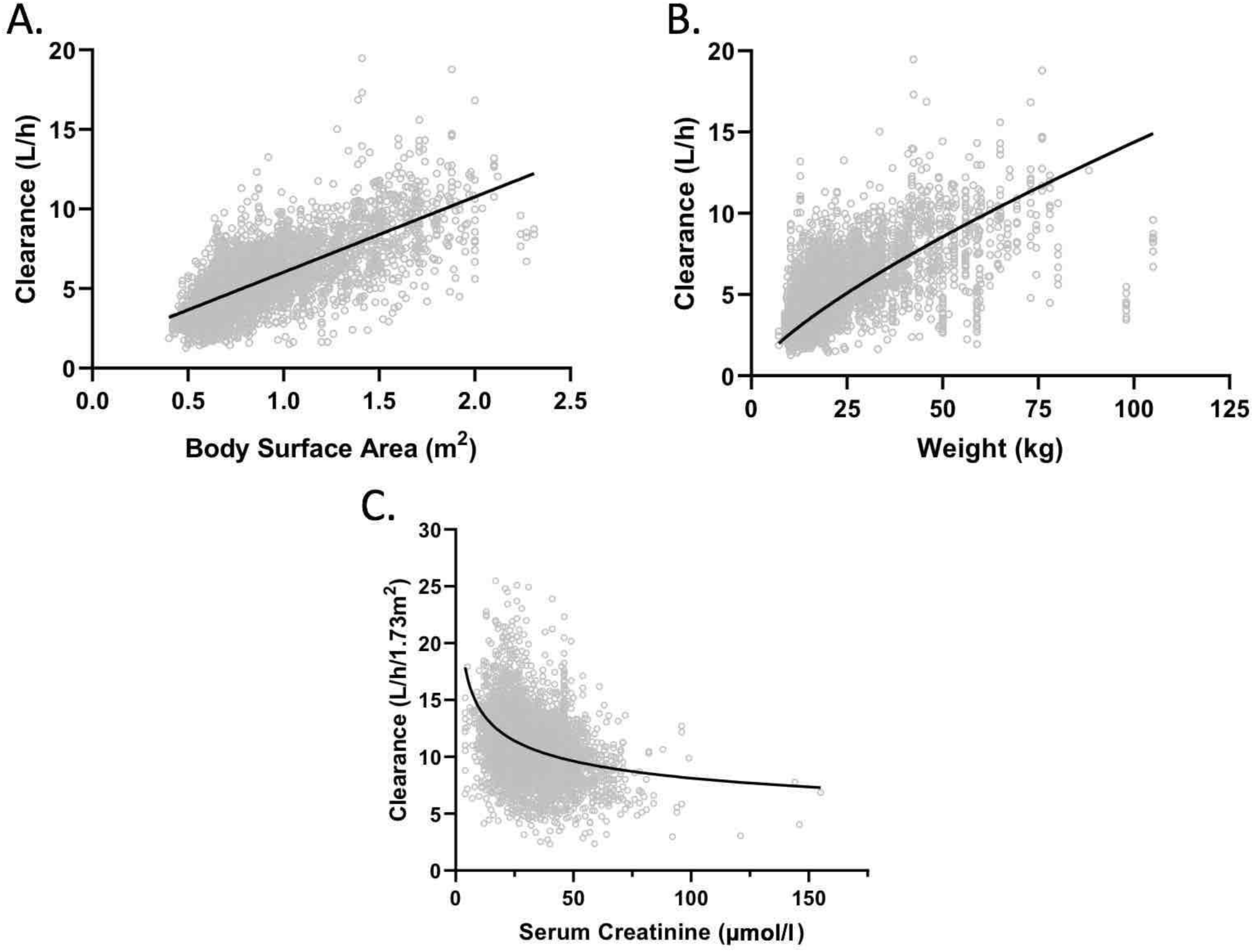
Covariate analysis. The coefficient of determination for BSA and the estimated clearance (A, R2 = 0.54) is a stronger relationship than the coefficient of determination for weight allometrically scaled to 70 kg and the estimated clearance (B, R2= 0.25). Serum creatinine demonstrated a non-linear with clearance normalized to BSA (C). The gray dots represent clearance estimates at the first serum creatinine level for each patient course.

The parameter estimates for the final model are presented in **Table 3**. Model estimates for the inter-individual variability of V1 and Q2 approached zero and were therefore fixed parameters (31, 32).

**Table 3:** Final model parameter estimates

| Parameters | Mean | Relative Standard Error (%) | Interindividual Variability | Relative Standard Error (%) |
| --- | --- | --- | --- | --- |
| CL<br>(L/h/1.73m<sup>2</sup>) | 11 | 0.7 | 0.08 | 4.7 |
| V1<br>(L/1.73m<sup>2</sup>) | 16.5 | 5.2 | 0 FIX | – |
| Q2<br>(L/h/1.73m<sup>2</sup>) | 0.602 | 4.1 | 0 FIX | – |
| V2<br>(L/1.73m<sup>2</sup>) | 4.55 | 3.4 | 0.12 | 4.3 |
| Q3<br>(L/h/1.73m<sup>2</sup>) | 0.111 | 2 | 0.13 | 6.7 |
| V3<br>(L/1.73m<sup>2</sup>) | 13.1 | 5 | 0.10 | 14.4 |
| SCR | -0.247 | 5.7 | – | – |

CL: Clearance of methotrexate from the central compartment
V1: Volume of distribution of methotrexate in the central compartment
Q2: Inter-compartmental clearance for vascular peripheral compartment
V2: Volume of distribution of methotrexate in the vascular compartment
Q3: Inter-compartmental clearance for the non-vascular compartment
V3: Volume of distribution of methotrexate in the non-vascular compartment
SCR: Pharmacokinetic estimate for serum creatinine

### Evaluation of MTX Population PK Models

Three models were implemented and tested as part of the MTXPK.org platform using the MTXSim prototype. Model parameters for each of the published population PK models are summarized in **Table S1, Supplementary Materials**. The model parameters were integrated into the MTXSim prototype with Edsim++ PK engine functionality (27, 28), which enabled us to load patient data from a previously published cohort (30) onto each model to generate individual Bayesian estimated PK estimations. Comparisons between PK estimates and observed values were made. The new three-compartment model demonstrated the least model bias and best model precision compared to other published models (**Table S2, Supplementary Materials**). The three-compartment model performed well among several validation cohorts that included all ages and indications (Table S3, Supplementary Materials).

### Web-based Clinical Decision Support Tool

The three-compartment model was selected as the default MTX population PK model to be integrated into the MTXPK.org webtool. This online, free-to-use tool allows the user to enter patient data that can be locally saved and reloaded later to add more information; however, nothing is stored by the website. The user will begin by entering the patient’s demographics, disease indication, and treatment regimen (**Figure 4A**) followed by the patient’s plasma MTX concentrations and SCr concentrations (**Figure 4B**). Once finished, the user can generate the individual’s personalized concentration-time curve, ‘Time to Elimination Threshold,’ and estimated area under the elimination curve (AUC) (**Figure 4C**). The user can immediately visually compare the patient’s elimination curve to that of the population average, the consensus guideline thresholds, and the 2 SD > mean elimination curve described in the glucarpidase label. The user is provided with quantitative metrics as well; when the cursor hovers over the concentration-time curve, estimated plasma MTX concentration values are provided with the population average and 2 SD estimates for the same time point. In the case of our example patient, which contains real PK data, the MTX concentration at 23 hours is higher than the actionable MTX concentrations outlined in the glucarpidase guideline and borderline on the glucarpidase drug label. Looking at the 36-hour time point, this patient is below the glucarpidase consensus guideline threshold, but remains higher than the glucarpidase label threshold. At hours 42 and 48, our example patient’s plasma concentrations equal that of the actionable concentrations outlined in the glucarpidase consensus guideline. Following the glucarpidase drug label and consensus guideline, our example patient would fit the criteria for glucarpidase administration. **Figure 4D** illustrates the full concentration-time curve of our example patient, which resembles that of the Bayesian estimated forecasting seen in **Figure 4C**. Furthermore, the user can also utilize the ‘Time to Elimination Threshold’ and estimated AUC located in the top of the window for additional guidance. Lastly, the user has the capability to generate and print the patient’s personalized PK report; this can be given to the patient’s family, stored physically in a patient’s file, or uploaded into the electronic health record for future reference.

**Figure 4:**
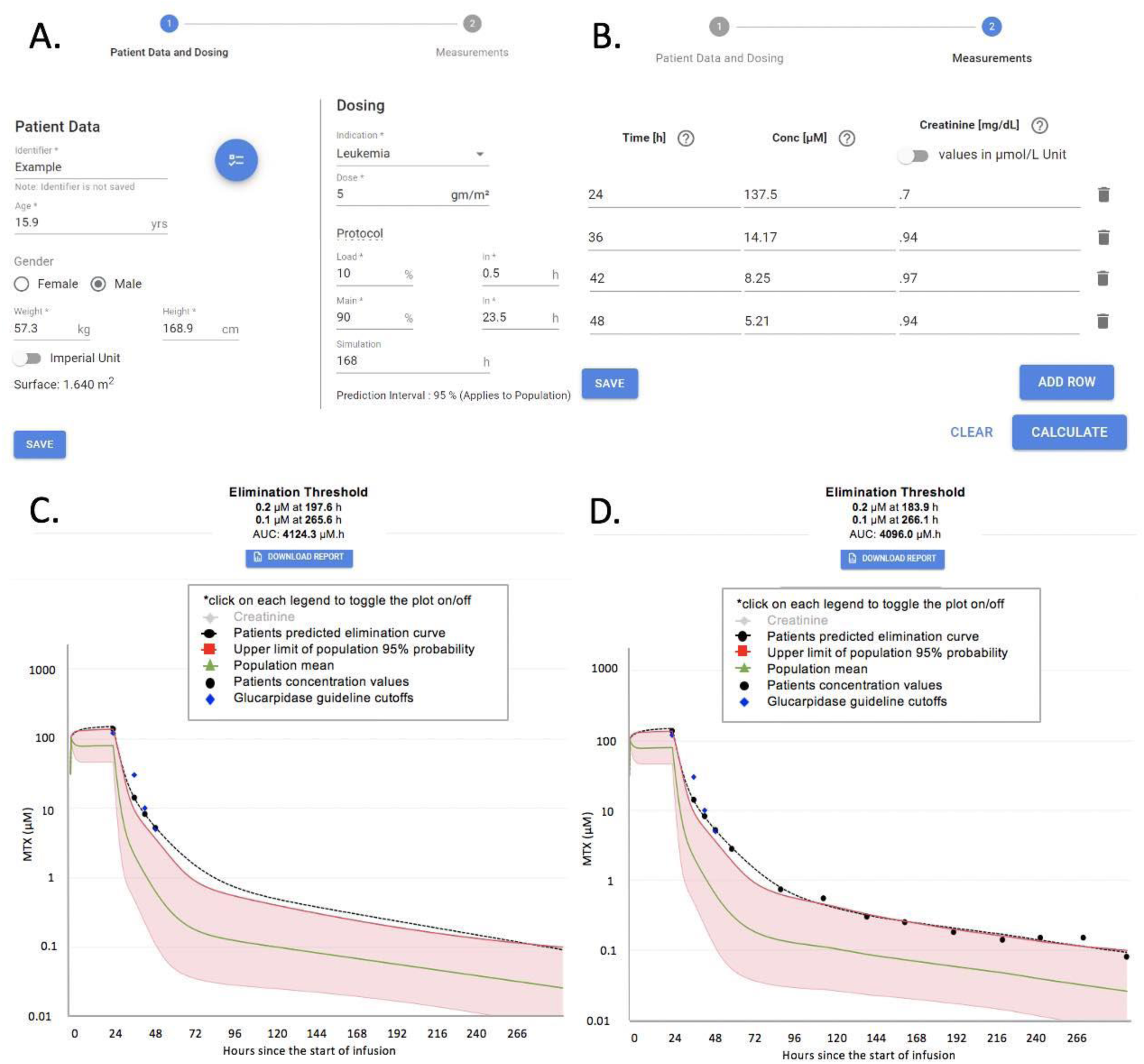
Screenshots of MTXPK.org. A, Patient demographics are easily loaded into the ‘Patient Data and Dosing’ screen. Plasma MTX concentrations and SCr levels are added to the ‘Measurements’ tab (B). After hitting ‘Calculate,’ the individualized PK estimates and concentration-time curve are generated. With the use of MTXPK.org, the user would conclude that this patient would meet the indication for glucarpidase since the individualized PK elimination curve (black line) is higher than the elimination curve for the population (green line), the 2 SDs (red line), and is close to the concentrations outline by the glucarpidase guideline (purple diamonds). The full concentration-time curve for the example patient (D) demonstrates the accurate forecasting of plasma MTX concentrations.

## Discussion

The purpose of this project was to develop a web-based clinical decision support tool that utilized a MTX population PK model and Bayesian estimation to facilitate optimal administration of glucarpidase for patients with delayed MTX clearance or high plasma MTX concentrations following an IV administration of HD MTX.

The current clinical challenges pertaining to optimal administration of glucarpidase arise when the time of a collected blood sample does not correspond to the time points outlined by the glucarpidase consensus guideline, appear very close to the consensus guideline’s actionable concentrations, or reveal rapidly rising SCr concentrations. MTXPK.org is able to mitigate these clinical challenges by facilitating model-informed administration of glucarpidase through the use of population PK modeling and a posteriori Bayesian estimation. The web-based clinical decision support tool works by allowing the user to enter the patient’s demographics, real-time drug concentrations, and SCr concentrations into the tool (**Figure 4A, B**). The tool then models the individual’s information and simulates a personalized elimination curve using Bayesian estimation (**Figure 4C**). MTXPK.org illustrates the average elimination curve for the population surrounded by the 2 SD > the mean elimination curve, described in the glucarpidase drug label (10), and the actionable plasma MTX concentrations at specified time points, outlined in the glucarpidase guideline (11). The clinician is provided with personalized visual and numeric information pertaining to their patient’s MTX elimination curve that can be compared to the glucarpidase drug label and consensus guideline to facilitate model-informed decisions about post-infusion supportive care and the administration of glucarpidase.

A similar tool to MTXPK.org was launched by Barrett et al. at Children’s Hospital of Philadelphia (CHOP) that integrated modeling and simulation into a hospital-based decision support tool to guide leucovorin rescue for patients receiving HD MTX (33).The Barrett MTX dashboard tool focused on the visualization of individualized elimination curves simulated from a posteriori Bayesian estimation of HD MTX. The elimination profile generated by the Barrett tool overlaid a leucovorin nomogram with additional dosing events relative to biomarkers of HD MTX toxicity presented to the clinician to guide dosing of MTX and rescue with leucovorin (34). Commercial and institutional support tools are available; however, MTXPK.org is the only free, publicly-available clinical decision support tool to inform post-infusion care following IV infusion of HD MTX.

In order to achieve optimal administration of glucarpidase, MTXPK.org needs to provide an accurate estimation of a patient’s individualized elimination curve and the population mean + 2 SD that corresponds to the glucarpidase label indication. The description of the average population elimination curve and associated 2 SD is captured in the default population PK model for MTXPK.org, which was developed using the extensive and PK-rich NOPHO database that included 5,535 plasma MTX concentrations collected > 96 hours from the start of infusion in 400 patients. Typical MTX population PK models are developed using PK data up to 44 hours after the start of infusion (18–23) or contain few samples from patients with delayed MTX clearance (29, 35, 36). These models overestimate MTX clearance and plasma concentrations when used to evaluate patients with delayed MTX clearance (as illustrated by the NOPHO two-compartment model in **Figure 2B)**. The three-compartment model provides a better estimation of the average elimination profile for the population and improved the description of the significant inter-patient variability compared to the two-compartment model, which are critical for the accurate visualization and appropriate interpretation of the glucarpidase drug label. Physiologically, the third compartment might be able to reflect the redistribution of intracellular MTX, which is the site of action (37, 38). Thus, MTXPK.org uses a well-defined population PK model to generate the visual and numeric information associated with the glucarpidase drug label and consensus guideline, which are used to facilitate the optimal administration of glucarpidase.

Aside from the accurate visualization of the glucarpidase label indication of 2 SD > the mean, MTXPK.org needs to accurately simulate a personalized elimination profile from the patient’s real-time drug concentrations and SCr concentrations using Bayesian estimation. The model parameters for the default population PK model for MTXPK.org serves as reliable, prior knowledge for a posteriori Bayesian estimation. This allows MTXPK.org to accurately forecast plasma MTX concentrations for a given patient using the information provided to the tool, with more drug concentrations shown to improve Bayesian estimations through improved Bayesian learning (34, 39, 40) (**Figure 4C, D**). The three-compartment model provides more accurate forecasting of plasma MTX concentrations following the infusion of HD MTX than the two compartment models we employed. Utilizing Bayesian estimation has the potential to limit the amount of blood samples collected during post-infusion care and thus reducing the cost of care. Accurate Bayesian estimation also means that MTXPK.org has the capability to estimate the total MTX exposure by calculating the AUC. Estimated MTX AUC provides an additional metric to govern therapy; maintaining a target exposure could guide therapeutic outcomes while mitigating patient toxicity. Along with an estimated AUC, MTXPK.org is able to estimate the ‘Time to Elimination Threshold’ in hours after the start of MTX infusion. This information can help establish expectations for appropriate inpatient duration and possibly reduce the financial costs to both the patient and institution (41). Most importantly, MTXPK.org utilizes a posteriori Bayesian estimation to illustrate a patient’s individualized elimination curve from real-time drug concentrations that can be visually and numerically assessed by the clinician to facilitate model-informed administration of glucarpidase.

Despite this model’s attractive performance characteristics, there are some limitations to the model. It is unclear whether the model fit to a primarily European patient population can be generalized to patients of non-European descent, although in other studies that included a more diverse cohort, race explained little of the inter-individual variability of MTX clearance (42). Additionally, while the NOPHO database proved to be the richest MTX PK data available to us, there were limited covariates to explore. There is considerable inter-individual variability in MTX clearance that previous publications have demonstrated to be partially explained by serum albumin (43), creatinine clearance (41, 44), estimated GFR (29), concomitant medications (45), and *SLCO1B1* genotype (18, 42, 46, 47). Furthermore, our PK data did not include information regarding aspects of supportive care, like fluid hydration and urine pH, which can impact MTX clearance (36, 48). Future work will aim to evaluate the clinical success of MTXPK.org and attempt to expand on covariates that may improve the model’s predictive performance. Moreover, incorporating plasma MTX samples from delayed MTX clearance patients after they have received glucarpidase into our default model will update MTXPK.org so that it can provide model-informed clinical care for these patients. Initial feedback from clinicians indicated that some would prefer it be included in the electronic health record to automatically pull in HD MTX dose, MTX and SCr measurements, similar to the CHOP clinical decision support tool. Clinicians practicing at hospitals without an electronic medical record have indicated that they would gladly enter the data on the webtool and use it on every patient receiving HD MTX. Thus far, MTXPK.org has been used by >900 unique users in at least 35 countries. As it stands, MTXPK.org is a free, web-based clinical decision support tool that can facilitate model-informed administration of glucarpidase for patients with delayed MTX clearance or high plasma MTX concentrations following the IV administration of HD MTX.

## Data Availability

Data may be available upon request

## Study Highlights

What is the current knowledge on the topic?

> Currently, both the glucarpidase drug label and glucarpidase consensus guideline are available to guide the clinical use of glucarpidase for patients with life-threatening MTX toxicity, but there are challenges to interpretation and implementation in the clinical setting.

What question did this study address?

> The study aimed to address the questions, “How can we facilitate the understanding of when glucarpidase is indicated for patients receiving HD MTX?” and “Can we develop a population PK model-informed decision platform that adequately describes the MTX PK in patients with delayed clearance?”

What does this study add to our knowledge?

> MTXPK.org, a free, online clinical decision support tool now exists to facilitate the understanding of when glucarpidase is indicated for patients receiving HD MTX.

How might this change clinical pharmacology or translational science?

> This tool will facilitate clinical decisions by clinical pharmacists and oncologists in when to administer glucarpidase, and provides an estimate of when the patient may reach the MTX concentration threshold for discharge.

## Acknowledgements

We are grateful to Maureen O’Brien and Jennifer Young for their guidance on the clinical utility of our decision support tool. We would like to thank all participating NOPHO institutions and physicians for their collection of MTX PK data used in this study. We appreciate Dr. Edmund Capparelli allowing us to test our model fit using data described in Kawakatsu et al, reference 30. Additionally, we are grateful to the patients and families that contributed their data to our study.

## Author Contributions

Z.L.T., T.M., N.C.P., B.B., A.N.S., W.S., N.F., A.A.V., J.H., & L.B.R. wrote the manuscript; Z.L.T., T.M., N.C.P., A.A.V., J.H., & L.B.R. performed research and analyzed data.

---

Supplementary:

**Figure S1:**
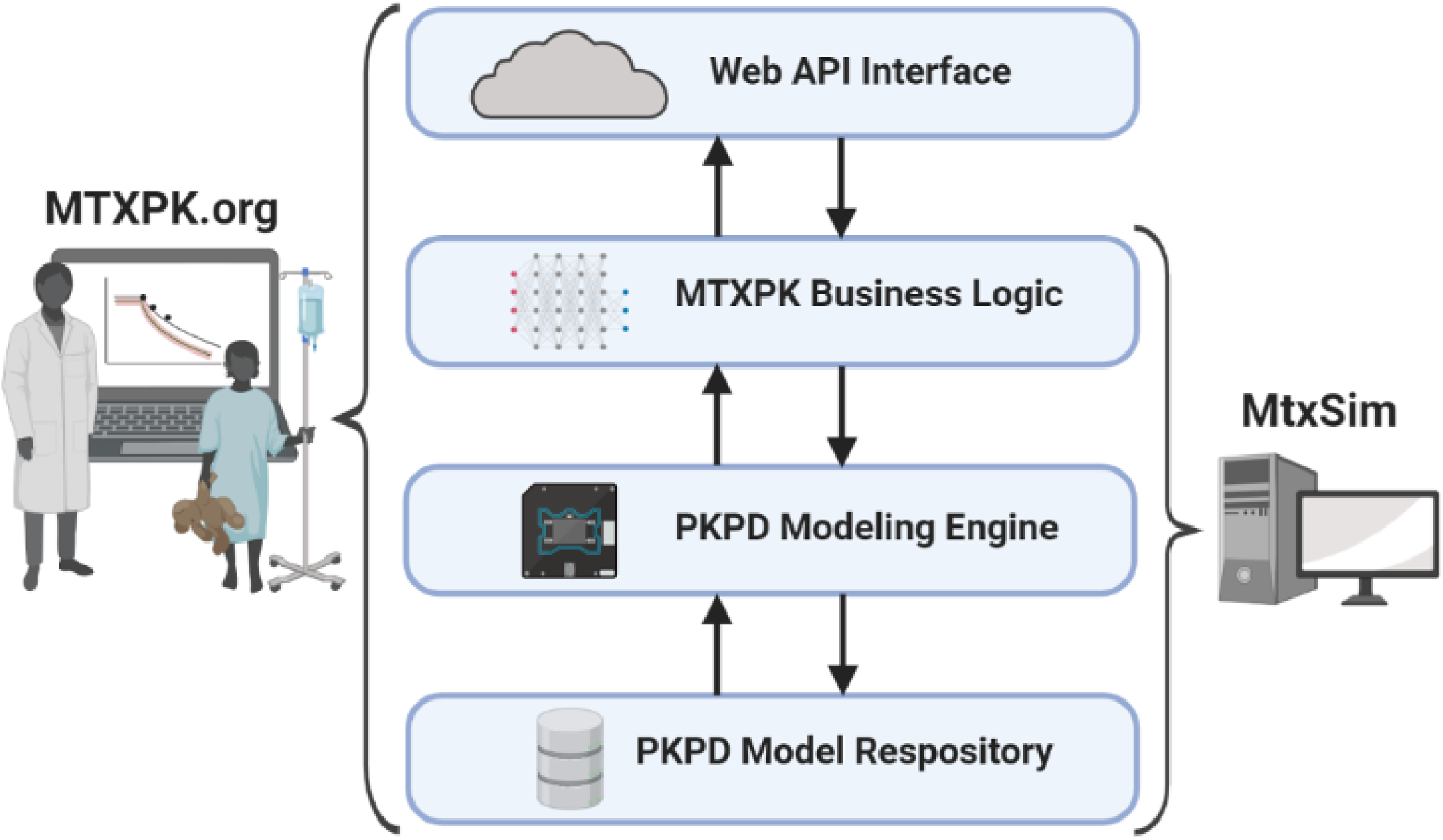
Architecture of MTXPK.org. MTXPK.org (left) is a web application that uses a Web API interface to interact with the end user. Commands are entered by the user then are automatically translated down a layer to the MTXPK business logic, where they undergo command processing. The logic string will call upon the general PKPD modeling engine, which uses a PKPD modeling repository to provide proper Bayesian estimation and simulation of the elimination curves. Processed information is sent back up the layering to the Web API to be presented to the end user. MtxSim (right) follows a similar logic string, but lacks the Web API interface to interact publicly with the end user. API: Application programming interface. PKPD: Pharmacokinetic/Pharmacodynamic.

**Figure S2:**
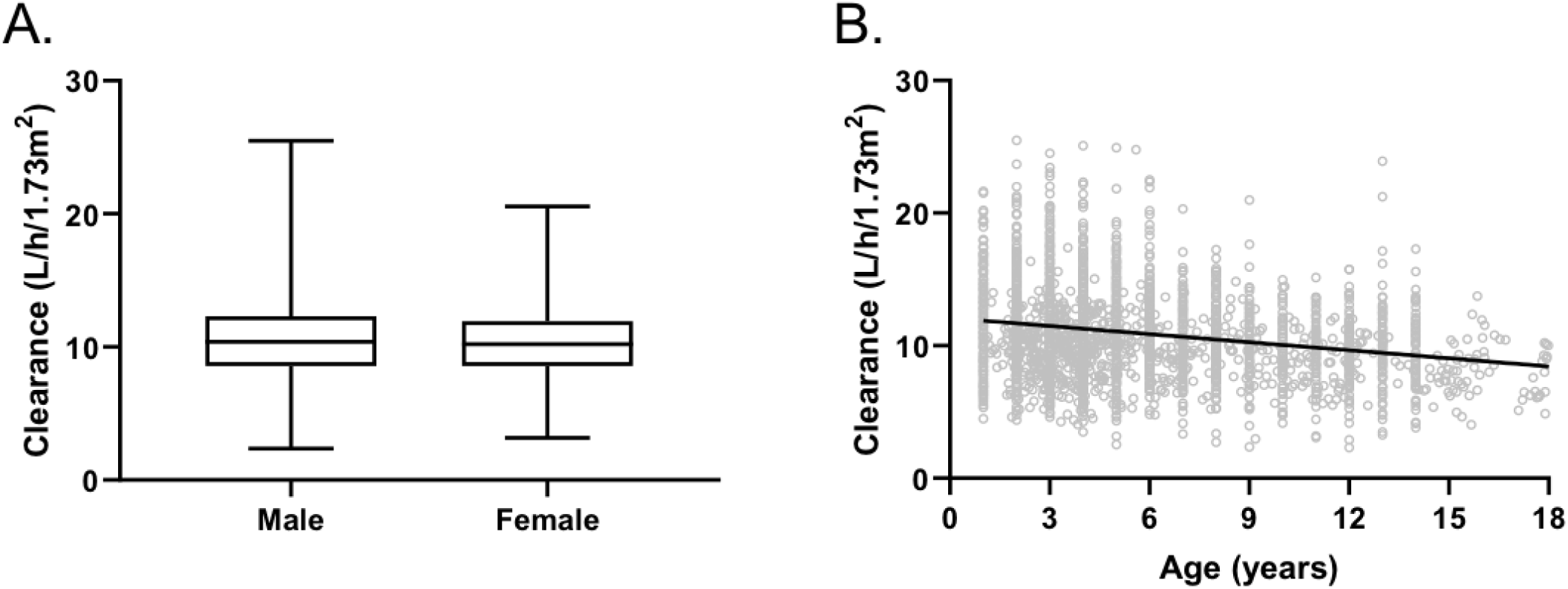
Additional covariate analysis. Once clearance was normalized to BSA, there was no significant effect of sex (A) on clearance estimates (p = 0.39). Age did not share a significant linear relationship with normalized clearance (B). The line within the box represents the median, the box represents the inter-quartile range, with the whiskers representing the minimum and maximum values. Gray circles depict the estimated clearance values at the given age at the start of treatment. The black line represents a linear regression.

**Figure S3:**
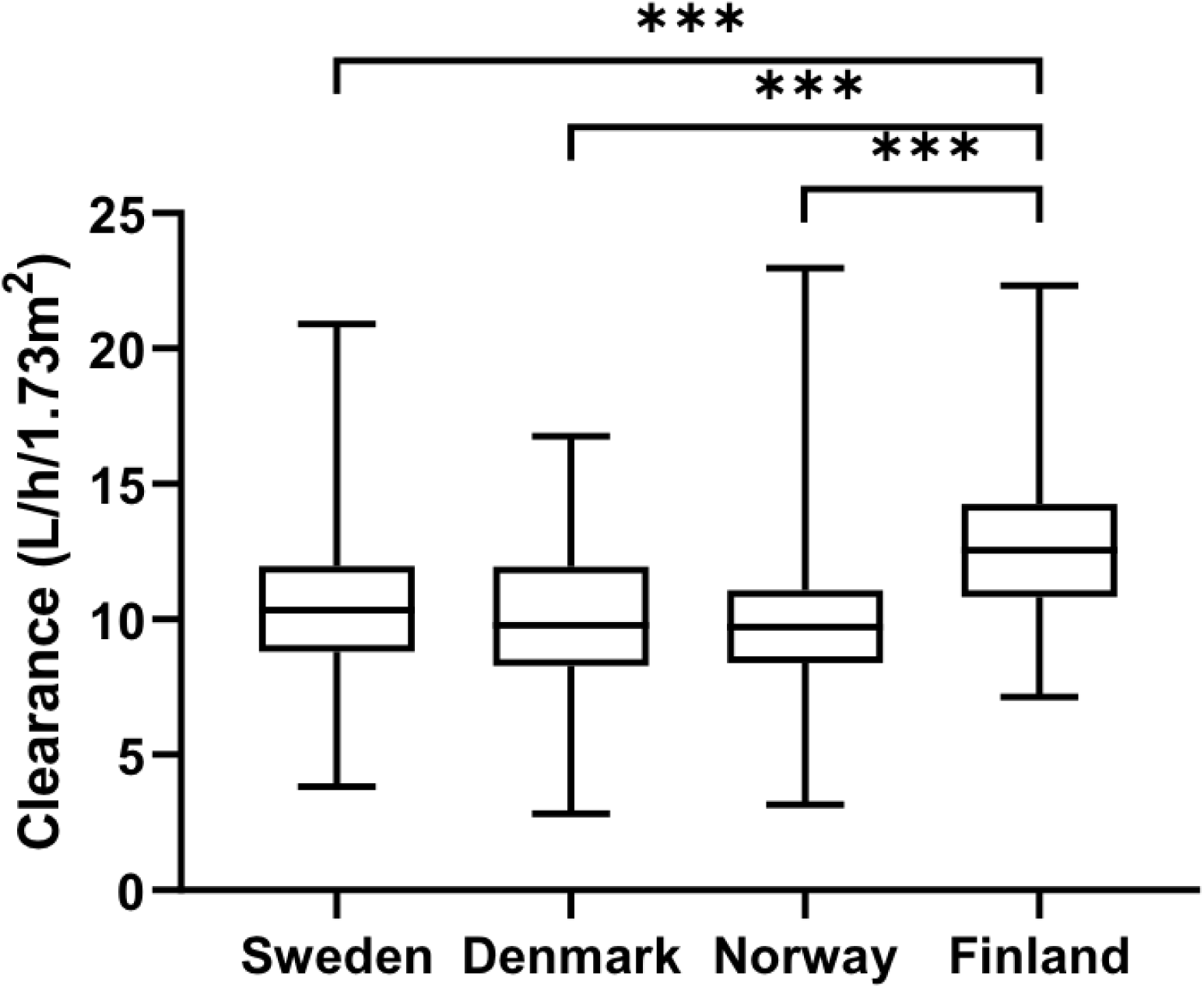
Impact of country of treatment on clearance estimates. Finnish patients had an estimated 26% faster clearance compared to Swedish, Danish, and Norwegian patients (p < 0.001). The line within the box represents the median value. The box represents the inter-quartile range with the whiskers representing the minimum and maximum values. *** corresponds to a p < 0.0001.

Supplementary:

**Table S1:** Model parameters for evaluation

| Model Name | Trevino <sup>a</sup> (18) |  | Kawakatsu <sup>ab</sup> (29) |  | NOPHO <sup>b</sup> - 3CP |  |
| --- | --- | --- | --- | --- | --- | --- |
| CL | 111.9 <sup>c</sup> | ml/min/m<sup>2</sup> | 12 | L/h/70kg | 11 | L/h/1.73m<sup>2</sup> |
|  | 10.9 <sup>de</sup> | L/h/1.73m<sup>2</sup> | 10.51 | L/h/1.73m<sup>2</sup> |  |  |
| V(1) | - | - | 52 | L/70kg | 16.5 | L/1.73m<sup>2</sup> |
|  | - | - | 45.54 | L/1.73m<sup>2</sup> |  |  |
| Q(2) | - | - | 0.1 | L/h/70kg | 0.602 | L/h/1.73m<sup>2</sup> |
|  | - | - | 0.087 | L/h/1.73m<sup>2</sup> |  |  |
| V2 | - | - | 5.6 | L/70kg | 4.55 | L/1.73m<sup>2</sup> |
|  | - | - | 4.9 | L/1.73m<sup>2</sup> |  |  |
| Q3 | - | - | - | - | 0.111 | L/h/1.73m<sup>2</sup> |
|  | - | - | - | - |  |  |
| V3 | - | - | - | - | 13.1 | L/1.73m<sup>2</sup> |
|  | - | - | - | - |  |  |
| Vc | 9 | L/m<sup>2</sup> | - | - | - | - |
| Ke | 0.7 | hour<sup>-1</sup> | - | - | - | - |
| Kcp | 0.08 | hour<sup>-1</sup> | - | - | - | - |
| Kpc | 0.11 | hour<sup>-1</sup> | - | - | - | - |
| Rationale | Two-compartment model used by St. Jude and mtz.stjude.org |  | Two-compartment model with eGFR included in the model |  | Novel three-compartment model using NOPHO PK data |  |

<sup>a</sup> PK parameters were estimated to a normalized 1.73m<sup>2</sup>
<sup>b</sup> Additional population PK model parameters exist for this model
<sup>c</sup> Value was pulled from the mtz.stjude.org website
<sup>d</sup> Calculated value derived from Ke and Vc
<sup>e</sup> 11.61 L/h/1.73m<sup>2</sup> would be the value if derived from mtz.stjude.org
CL: Clearance of methotrexate from the central compartment
V1: Volume of distribution of methotrexate in the central compartment
V2: Volume of distribution of methotrexate in the vascular compartment
Q3: Inter-compartmental clearance for the non-vascular compartment
V3: Volume of distribution of methotrexate in the non-vascular compartment
Ke: Elimination rate constant from the central compartment
Kcp: Elimination rate constant from the central compartment to peripheral compartment
Kpc: Elimination rate constant from the peripheral compartment to central compartment
CP: Compartment

**Table S2:** Evaluations of MTX population PK models

| Model | Population |  | Individual |  |
| --- | --- | --- | --- | --- |
|  | Bias (%) | Prec (%) | Bias (%) | Prec (%) |
| **Trevino et al. (18)** | -38 | 87 | 0.3 | 20 |
| **Kawakatsu et al. (29)** | 7 | 60 | 0.4 | 12 |
| **NOPHO (3-CP)** | -15 | 55 | -0.2 | 9 |

Prec: Precision
CP: Compartment

**Table S3:**
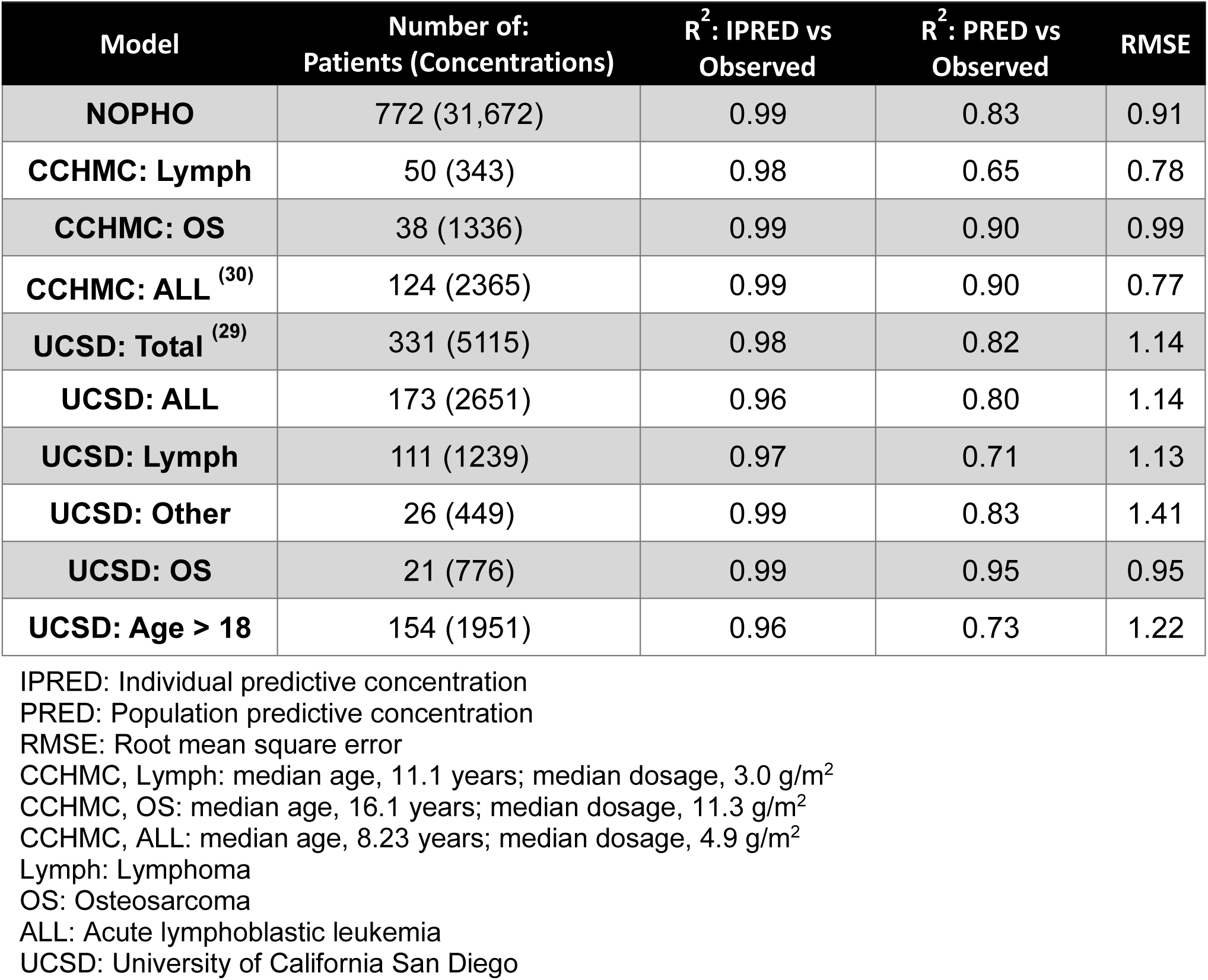
External validation of MTXPK.org tool

| Model | Number of:<br>Patients (Concentrations) | R<sup>2</sup>: IPRED vs<br>Observed | R<sup>2</sup>: PRED vs<br>Observed | RMSE |
| --- | --- | --- | --- | --- |
| **NOPHO** | 772 (31,672) | 0.99 | 0.83 | 0.91 |
| **CCHMC: Lymph** | 50 (343) | 0.98 | 0.65 | 0.78 |
| **CCHMC: OS** | 38 (1336) | 0.99 | 0.90 | 0.99 |
| **CCHMC: ALL** <sup>(30)</sup> | 124 (2365) | 0.99 | 0.90 | 0.77 |
| **UCSD: Total** <sup>(29)</sup> | 331 (5115) | 0.98 | 0.82 | 1.14 |
| **UCSD: ALL** | 173 (2651) | 0.96 | 0.80 | 1.14 |
| **UCSD: Lymph** | 111 (1239) | 0.97 | 0.71 | 1.13 |
| **UCSD: Other** | 26 (449) | 0.99 | 0.83 | 1.41 |
| **UCSD: OS** | 21 (776) | 0.99 | 0.95 | 0.95 |
| **UCSD: Age > 18** | 154 (1951) | 0.96 | 0.73 | 1.22 |

IPRED: Individual predictive concentration
PRED: Population predictive concentration
RMSE: Root mean square error
CCHMC, Lymph: median age, 11.1 years; median dosage, 3.0 g/m<sup>2</sup>
CCHMC, OS: median age, 16.1 years; median dosage, 11.3 g/m<sup>2</sup>
CCHMC, ALL: median age, 8.23 years; median dosage, 4.9 g/m<sup>2</sup>
Lymph: Lymphoma
OS: Osteosarcoma
ALL: Acute lymphoblastic leukemia
UCSD: University of California San Diego

## Supplemental Methods

### Model Development

We performed a population PK analysis of HD MTX using nonlinear mixed-effects modeling implemented in NONMEM^®^ version 7.2.0 (ICON, Ellicott City, MD) and validated using KinPop++. Two- and three-compartment structural models with either an additive residual error model, proportional residual error model, or combined additive and proportional error model were considered. The FOCE method with interaction was used. The three-compartment with an exponential residual error model was chosen as the base model after considering objective function value (OFV) and goodness-of-fit (GOF) plots. Clearance in L/h was the PK parameter of interest. The final model included additional PK parameters: volume of distribution of the central compartment, V1, in L; the inter-compartmental clearance for vascular peripheral compartment, Q2, L/h; the volume of distribution of the vascular peripheral compartment, V2, L; the inter-compartmental clearance for the non-vascular peripheral compartment, Q3, L/h; and the volume of distribution of the non-vascular peripheral compartment. Forty-eight patients and 679 concentrations were excluded from the PK analysis due to missing dosing information and an additional 753 MTX concentrations were considered implausible by the investigators and were excluded in the NONMEM analysis. Validation of the base model was achieved using KinPop++ and all 33,104 MTX concentrations. The final base model included between-subject variance components for the each of the PK parameters previously listed (Table 3)

### Covariate Analysis

Clinical (SCr) and demographic (BSA, weight, age, sex) data collected during the course of the MTX treatment were included in the covariate analysis. A stepwise covariate modeling procedure was implemented using the three-compartment structural model (1). The stepwise inclusion of a covariate is based on a drop in NONMEM OFV > 3.84 (p < 0.05) (2, 3). All clinical and demographic covariates, except for country, were included in the covariate analysis. The effect of country on MTX clearance was analyzed using a one-way ANOVA after the final model was developed. Finnish patients are said to receive their hydration with bicarbonate approximately 12 hours before the initiation of HD MTX compared to 4 hours for the remaining Baltic and Nordic patients. Effects of pre-hydration on MTX PK have been published (4), but was not included in our data file. SCr as a time-varying covariate. GOF plots were generated following a significant reduction in OFV. Observed MTX concentrations were plotted against the individual (IPRED) and population (PRED) predicted MTX concentrations and compared to the line of identity. The conditional weighted residuals (CWRES) were analyzed against the PRED and time after infusion to determine if there was significant non-zero bias in the model.

### Bayesian Estimation

A module for Bayesian estimation was included in the MTXPK.org platform using a three-compartment structure model developed using the visual PKPD model designer Edsim++ (Mediware, Prague, Czech Republic) and executed using an Edsim++ compatible PKPD modeling engine (5, 6). The curve-fitting capability allows individual PK parameter estimation using a Bayesian procedure (7). Data used for the model-informed individual PK estimation included the patient demographics, dose administered, infusion duration, MTX plasma concentrations, and SCr levels. The individualized elimination curve allows for immediate quantitative and visual feedback on a patient’s MTX clearance in reference to the average concentration-time curve for the population, which is generated from the final model population parameters with ± 2 SD graphically indicated by a shaded area (**Figure 4C**). The upper-boundary represents the indication for glucarpidase, where the MTX clearance is >2 SDs below the mean for the given dose (8). The confidence interval calculation is based on the Delta method (9), which employs the basic laws of error propagation. The actual implementation is described in detail by Proost (10) and implemented in PK software packages like MW/Pharm (11).

